# Obesogenic Assesment of the Primary Schools Environment: A Qualitative Narrative Analysis

**DOI:** 10.1101/2020.12.02.20243014

**Authors:** Siti Rohaiza Ahmad, Robert Bush

## Abstract

**Background:** While multiple influences on children’s food choices and eating habits have been proposed, including genetic, familial and environmental influences, it is increasingly the environments they live in, including schools, that are receiving attention as sites for understanding health-related behaviors.

**Aim:** To analyse the infrastructure of Brunei’s primary schools in terms of the level of obesogenicity and how the condition can be improved for the future.

**Methods:** Site of research was five purposely selected primary schools in Brunei Darussalam. In this research, the obesogenic audit tool used were developed from the Primary School Environmental Audit (PSEA) tool.

**Results:** Similarities and differences to the nutrition and physical activity environment across the five different schools was found. Consistency was observed across all school sites in the MOE and MOH formal rules and regulations that were applied to the school food-services operations.

**Conclusion:** The use of a modified PSEA audit tool, has successfully enabled the successful identification of the various elements of primary schools in Brunei that may have contributed to an obesogenic environment. The research approach in this study was to not only examine a school’s internal environment but its external environment as well. This ensured a more complete overview of what was in-place, allowing assessment of what could be better improved to make it more conducive to childhood obesity prevention.

## INTRODUCTION

Childhood is a period of critical development and growth, dependent in part on adequate nutrition and physical activity to ensure health and wellbeing (Hay, 2005; Huston, 2006). While multiple influences on children’s food choices and eating habits have been proposed, including genetic, familial and environmental influences (Scaglioni, Arrizza, Vecchi, & Tedeschi, 2011), it is increasingly the environments they live in, including schools, that are receiving attention as sites for understanding health-related behaviours (Birch & Davison, 2001; Kral & Rauh, 2010), and for public health intervention research and policy (Kirk, Penney, & McHugh, 2010).

Providing good childhood nutrition and encouraging good eating habits is challenging, given an environment of competing food messages, yet remains a critical task in guiding children along the passage to healthy adulthood (Bannon & Schwartz, 2006). The global food industry has seen rapid expansion in recent decades with increasing control of the food markets, and diversification of products (World Health Organization, 2013). Along with these changes, an expansion of energy dense nutrient poor (EDNP) foods and beverages has become available to consumers, including child consumers. Evidence has shown that high intake of EDNP foods and beverages including soft drinks, chips, biscuits and confectionaries have been associated with the increased likelihood of childhood obesity (World Health Organization, 2013). Furthermore, the increase in a sedentary lifestyle has also led to increased incidences of energy balance, thus promoting weight gain (Carlson, Crespo, Sallis, Patterson & Elder, 2012; World Health Organization, 2013).

Childhood obesity is rapidly increasing across the globe (Karnik & Kanekar, 2015; Fleming, et al., 2014). The International Obesity Task Force (IOTF) has reported that about 10% of school-aged children (5 to 17 years old) globally are found to be in the overweight category. A quarter out of the 10% are in the obese category (Lobstein, Baur, Uauy, & TaskForce, 2004). Asian countries, including Brunei, are also facing increasing childhood obesity cases (Ministry of Health, Brunei 2011). The prevalence of childhood obesity monitored among those in primary school Year 1, 4, and 6 and secondary Year 2 shows an increasing trend from 11.8% in 2005 to 18% in 2014. Health problems caused by childhood obesity are likely to persist into adulthood (Lloyd, Langley-Evans, & McMullen, 2012). The consequences of obesity are detrimental to the country’s economy, need to provide medical specialists and other hospital services will increase substantially (Selassie & Sinha, 2011; Withrow & Alter, 2011).

School setting is an ideal place to influence health, because children are exposed to the school environment in the first two decade of their life. Schools environment offers continuous and intensive contact for children outside the home environment (Carter & Swinburn, 2004). School is mostly compulsory for all children across the globe. In Brunei, the local policy is to provide a minimum of 12 years of education (7 years primary education and 6 years secondary education) (Ministry of Education, Brunei 2014). Therefore, continuous and intensive contacts made school an ideal setting for influencing health, including obesity prevention programs.

In this research, the primary school food and physical activity environments are investigated. The aim of the research is to analyze the infrastructure of Brunei’s primary schools in terms of the level of obesogenicity and how the condition can be improved for the future.

## METHOD

### Site of Research

The site of the research is Brunei Darussalam. Brunei is located in South-East Asia, on the northwest coast of the island of Borneo. Five primary schools were selected. Initially, the schools were selected based on their geographical location and their environment to reflect the different areas of living in Brunei. The selection of the schools was via purposeful sampling designed to identify a cross-section of schools.

### Audit Tool Development

In this research, the audit tool used were developed from the Primary School Environmental Audit (PSEA) tool (Glass & McAtee, 2006).. The full detail of the audit has been reported in a separate publication (Ahmad et al., 2016). In this research, areas covered different dimensions of the school environment which includes analysis of the ‘internal canteen service’, ‘external canteen service/food stall vendors’, ‘school food/nutrition policy’, ‘nutrition environment’, ‘school physical activity policy’, ‘school physical activity environment’ and ‘external neighborhood physical activity environment’. The questionnaires were completed by the researcher (SRA) over a three-day site visit to each school, making observations and seeking specific information from the various school community members.

The schools involved in the study were provided with feedback on the analysis of their environment. The results of the analysis were delivered to the school community via a feedback forum or/and discussion group. This is a form of member-checking to check if the data interpretation could be verified by the school stakeholders (Collingridge & Gantt, 2008). It therefore also provided the opportunity to identify any potential missed information about the schools that should be included in the analysis. In summary (Figure 1), the steps of the data collections were data collection, interpretation and preparation of summary document for the school, presentation or report back to the school and lastly, further discussion with the school stakeholders (Ahmad et al., 2016).

**Figure 1.**
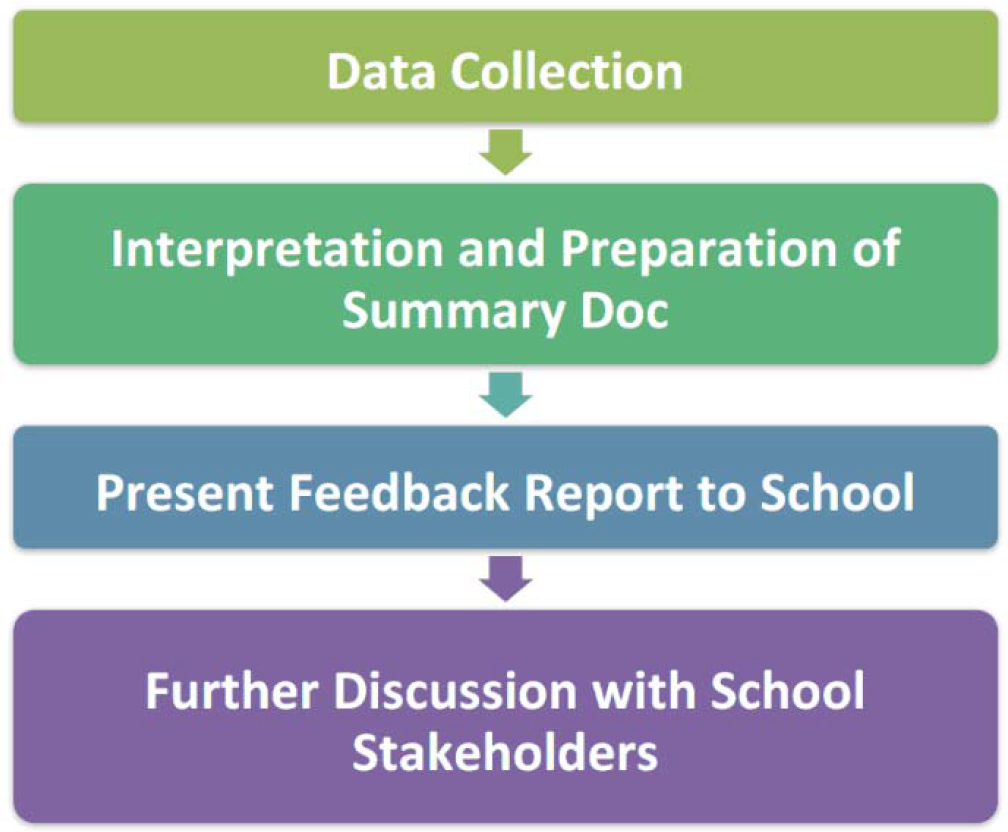
Step by step for conducting the school audit (Ahmad et al., 2016)

## RESULTS

### School Demographic

Table 1 shows the descriptions of the primary school case-study settings. The student-to-teacher ratio for the schools varied between 9:1 and 13:1. School C has the highest student-to-teacher ratio compared with the other schools. In terms of the building structure, all of the schools were government owned schools, with similar building structures, including the canteen building structures and other facilities, observed across the school sites

**Table 1.** Descriptions of the primary school case-study settings.

| **School** | **Location/mini descriptions about the school** | **Date the initial audit/assessments were conducted** | **Number of students and teachers (student to teacher ratio)** | **Buildings** |
| --- | --- | --- | --- | --- |
| **A** | A small village located in a slightly hilly area, with some food stalls selling cheap foods around the school area, some convenience stores and restaurants. | 15 to 17 October 2012 | 219 students and 24 teachers (9:1) | Two blocks for classrooms, one block for administration and library, one sports hall, and one small school canteen |
| **B** | Large residential area, with many food stall vendors near the school gate. | 14 to 17 January 2013 | 516 students and 45 teachers (11:1) | Two blocks for classrooms, one block for administration and library, one sports hall, and one small school canteen |
| **C** | Oil rich district of Brunei. | 27 to 29 February 2013 | 461 students and 34 teachers (13:1) | Two blocks for classrooms, one block for administration, one sports hall, and one small school canteen |
| **D** | Remote area of Brunei (45 minutes by boat from the main city). This district is still under-developed because of preservation of many of the forests in the area. | 30 April and 2 May 2013 | 135 students and 15 teachers (9:1) | Two blocks for classrooms, one block for administration, one sports hall, and one small school canteen |
| **E** | A water village, with many cheap food stalls located outside the school gate. The water village reflects one of the oldest and well-preserved local heritages of Brunei. | 13 and 15 May 2013 | 265 students and 31 teachers (9:1) | Two blocks of classrooms with a small built-in school canteen and kitchen area, a sports hall, and an administration office |

### Nutrition environment: Similarities across the five schools

Consistency was observed across all school sites in the MOE and MOH formal rules and regulations that were applied to the school food-services operations. All of the five schools had one internal canteen service that opens every school day. These services were mainly operated by local Bruneians who run the canteens as for-profit business venture and no volunteer staff members are used. The canteen operators rent the canteen facilities from the school administration, based on the total number of students in the school (more students mean higher rental fees). In no instance was the profit from the school canteen reinjected into the school.

The hours of the normal school day for the morning session are from 7.00 am until 12.30 pm. All of the school sessions selected for this research were morning sessions. Hours of canteen operation are from 10.00 am until 10.30 am. School starts early during the day, however, no breakfast service is available for the students. Their break time is around morning tea time (10.00 to 10.30 am). No snack break was available from morning until the break time. Therefore, the break time is the only opportunity for students to eat in the morning. At the school canteen, the students normally select and purchase the available foods and beverages (i.e. no pre-order is available from the canteens for the students). Some students also bring food and beverages from home. In most of the schools, tables and chairs were prepared for the students to use when eating their meals. Sometimes, students bring their food and eat it inside their classroom or just stand outside their classroom while talking (a common sight for girls). There was no evidence of enforced rules regarding where the students can eat in the school grounds.

The external vendors normally operate immediately prior to the commencement of the school day, throughout the day, and at closing time; thus directly targeting students. Students were strictly prohibited by all of the five school administrations from going out of the school compound during school hours to buy from the food stall vendors. Despite this ban, the vendors continue to operate their business because the students make purchases after school hour (which is beyond the control of the school administration). The teachers are encouraged to make healthy choices when purchasing from the canteen, so that they represent good role models for healthy eating behaviours for students. The teachers buy from the school canteen and not from any external vendors to set a good example.

Generally all small packed food, such as packed sausages, nuggets and chips in cups, were available at a low cost (BND$0.50). Larger sized prepared-meals, such as rice served with chicken, commonly sold for BND$1. The prices for beverages range from BND$0.50 to BND$1. They reported that the most popular menu items are rice served with beef or chicken and finger foods, such as chicken nuggets, sausages and chips, predominantly meals prepared by the canteen operator in-house. The cheapest beverage price was BND$0.20 per serve, which was for a small iced Milo ™ sealed in plastic. In all of the schools, there was a feedback system whereby students and teachers could raise concerns about canteen item pricing with the canteen operator.

To ensure that canteen operators adhere to the Healthy Canteen Guidelines produced by the MOE, school canteens are monitored by the internal school administration including the principal, vice principal and the school’s canteen committee consisting of appointed teachers. No evidence was available that these committees met formally. One of the responsibilities of the canteen committee was monitoring of standards at the canteen as well as monitoring food brought from home. The frequency of monitoring of the school canteen, as well as monitoring of food brought from home by the student varies, with committee members at some schools being more vigilant than others. During the canteen inspections, the school administration observes whether the food and drinks sold adhere to the Healthy Canteen Guidelines, as well as the level of cleanliness, for the purpose of monitoring of standards and compliance with the Guidelines. For School E, the principal considered the canteen operator to be ‘*very cooperative*’ in adhering to the Healthy Canteen Guidelines, and thus reduced the canteen checks to once every couple of months. For School B, sweet chocolate cake (chocolate powder based) was sold ‘secretly’ (hidden under the counter requiring students to ask for it) to the students and teachers. Bottled plain water (600 ml) and fruit tea packet drinks were the only beverages sold. Mango drink and soya bean drink were previously available, but they were discontinued as they represented low profit items for the canteen.

Although all school canteen operators are required to adhere to the Healthy Canteen Guidelines, there were incidences of some school canteen operators failing to follow these guidelines. It is highly likely that this was due to some instructions being misinterpreted. Another possibility is that canteen operators believed that there were probably no consequences for ignoring the guidelines. For example, the guidelines state that frying (with a small amount of oil only, to prevent food sticking on the hot plate) is allowed a maximum of two times per week. However, the term ‘frying’ has been referred to as deep frying by a canteen operator in School D. In most cases, menu selections were based on easy preparation and profitability. School A and School E had canteen operators who emphasised the importance of healthy cooking methods (using a small amount of oil, steaming or boiling) and menus in their canteen. School A uses the air-fryer to prepare some of their foods. Canteens at School A, School B, School D and School E sold cut-up fruits either daily or a couple of days per week. All of the canteen operators admitted that fruit sales were quite poor, but some canteen operator still sells them despite the low demand.

Some primary school subjects including science, Malay, and civics have the potential for their curriculum to cover content related to nutrition. However, in the absence of any attempt from MOE to ensure that this occurs, this is currently entirely dependent on individual schools and/or teachers’ creativity and motivation to do this. One of the schools introduced staff development activities on every Wednesday afternoon related to healthy food preparation and cooking methods to increase teachers’ awareness.

Teachers in the schools also had their own eating routines and traditions. For example, some teachers bring any outside food and also soft drinks to school but hide them inside the fridge to be consumed during lunch time when the students have already left for home. None of the teachers on the school canteen committee were found to have qualifications in nutrition or any training related to nutrition.

There are also other similarities which include restrictions on students buying from external food stall vendors during school hour. However, any time after the school hours, the school cannot stop the students from buying from the external food stall vendors. Posters related to healthy eating and physical activity are displayed around the school compounds, and many of these posters were obtained by the school principals from the MOH. School A also has posters designed and made by students in the dining area near the school canteen; some students do read them during break time. None of the schools were found to have vending machines.

### Nutrition environment: Differences across the five schools

In terms of the external school environment, School A, School B and School E have unlicensed food stall vendors. School C and School D’s principal stated that any mobile vendors who located themselves in close proximity to the school compound were asked to move. The interpretation of ‘close proximity’ varied between schools; With School A and School E, external vendors were located immediately adjacent to the main school gate. With School B, as explained by the school principal, the vendors had previously been in the practice of locating themselves adjacent to the school gate, but have responded to requests by the school principal to move further away. These vendors sold processed packaged foods, although most of the food and beverages available for sale were home prepared, offering a range of colourful, hot and cold food and beverages to attract the attention of customers. These offerings included many choices high in fats, sugars and/or salt. These vendors supply varieties of food and drink that specifically appeal to primary school students. Compared with the school canteen, these vendors sold a greater variety of food and drinks that were less expensive than comparable items sold in the school canteens. The prohibition that restricted students’ use of external food stalls during school hours did not extend to teachers, who purchased from these food stall vendors during school hours. The school administration from School B and School E believes that the Municipal Department is the one that should be responsible for dealing with the issue of the food stall vendors. In one case, School E, the head villager was informed about the issue of food stall vendors, but there was reluctance to take any action because the owners of the stalls were mainly villagers who wanted to support their families.

In terms of the promotion of a healthy eating culture in each school, this relied on the enthusiasm of the school administration at each location. In some cases, academic achievement is seen as the school’s primary responsibility and main focus. In these instances, the promotion of healthy eating is seen as the responsibility of the MOH. In other cases, school administrators have introduced their own informal rules and guidelines, for example allowing only healthy food and drinks to be brought from home. Random checks of food and drinks brought from home were also conducted during school hours. For School A, during special events, such as sports day with parent’s involved, special guidelines were given by the school administration about the type of food and beverages, either for sale or self-consumed, that can be brought to the school. At another school, the administration did not allow a fast food restaurant to promote themselves by conducting activities in the school.

School D is the only school that has the School Lunch Programme running for 100% of their students (after 12.30 pm), which mainly provides healthy food for the students (the menu comes from MOE). The students were monitored during the program, to ensure they finished all their meals including the vegetables. However, the teachers admitted there was a wastage issue because the students didn’t really like some of the menu items. The principal also encourages teachers not to bring unhealthy food and drink to school.

### Physical activity: *Similarities across the five schools*

Formal physical education was implemented in all of the schools according to guidelines provided by the MOE. Time spent on the formal PA curriculum was 30 minutes per week for lower primary (Year 1 to 3) and 60 minutes per week for upper primary (Year 4 to 6). One hour was allocated every Saturday morning for an aerobic session for the teachers and students. Teachers had shown great enthusiasm in promoting physical activity among the students, despite the common issue of lack of proper sports facilities.

All of the schools conducted extracurricular activities (club activities) every week and some of the activities were related to physical activity. Students were also engaged in free play during the break time. Some sport facilities, mainly for badminton and netball, were provided for the students during break time by the teachers. However, not all of the schools practice this due to not having enough staff members to supervise the students during play. In most of the schools, students were also allowed to bring their own sports equipment at their own responsibility. The students were also encouraged to participate in sport competitions internally and externally.

Several elements that promote active travel were also looked at, including the ability to walk and cycle in the school neighbourhoods. For School B, pedestrian paths leading to the school were provided; however, it didn’t go as far as the housing catchment areas. For most of the schools, no proper pedestrian path was available (School A and School C). Zebra crossings were also provided outside School A and School B; however, based on my observation, some parents and students did not use them when crossing the road. In some of the schools, as part of an effort to strengthen safety, a teacher is appointed every day to oversee the students when their parents pick them up to go home (actively organised in School A – they have a schedule for the teacher).

It was unusual to find parents willing to allow their children to walk or cycle to and from school. The reasons given were because of the travel distance and the absence of safe and dedicated pedestrian or bike paths. Even though zebra crossings were fairly commonly sighted outside school grounds, there was little evidence of any other traffic calming measures: zebra crossings were not patrolled, and there were no school-zone speed limits in operation or signboards to remind drivers to drive slowly in the school neighbourhoods.

### Physical activity: Differences across the five schools

There were no formal departmental guidelines relating to physical activity in the schools. Some teachers in the schools have designed informal rules and guidelines related to physical activity for the students. The school administration and teachers made an effort to prepare suitable areas for sports with whatever facilities and creativity that they have. For example, some ground areas within the school compound were painted with chequered patterns and numbers for playing hop scotch (School B has this). Observations during the study indicated that students engaged in and enjoyed these activities when provided. However, safety is a concern when playing outside due to hot weather and uneven cemented ground. Another concern raised by the physical education teacher in one of the school was that some subject teachers perceived physical education as not important because these teachers always let the students off late to go to physical education classes. School C implemented a walk-to-school program for students living nearby so that they could walk to school together. For School C, because it is located on a water village, most of the students walk to school via the jetty or water taxi, but safety could be improved by installing fences.

The physical activities conducted across the five schools also vary, particularly during the school break time. In School A, students have the opportunity to play badminton and have free play within the school grounds or in the sports hall. School B has a special play area for the students, with lines prepared by the teachers for netball, football and hop scotch. This area was located very close to the teachers’ staffroom for easy supervision, even though no actual supervision was observed. However, the school principal does walk around the school compound every now and then to check on the activities of students.

School B has a playground, provided by the Brunei Government; however, due to poor maintenance (loose screws and broken parts), the playground cannot be used. For School C, the students were mainly engaged in free play during the break time. During break time at School E, teachers prepare a special game for the students called the gutter board game (Introduced by the Canadian English teacher). ‘Boat racer’ games were unique to School E: the students build a small boat from a piece of paper and slide them on the tiles, the boat that slides the furthest is deemed the winner. The boat racer game involves some physical activity, as students run to gain momentum so that their boat will slide further.

The physical education (PE) teachers were asked about the adequacy of the indoor and outdoor sports facilities and playgrounds in the school, as they were best placed to know how physical activity (PA) programs were conducted and whether space, equipment and other facilities were an issue when running PE lessons and during free play. PE teachers also look after the sports equipment so they are familiar with issues around the adequacy of the equipment. School B’s PE teacher was the only PE teacher who believed that she had adequate space for indoor PE classes or activities. All the schools’ PE teachers said that their outdoor space for physical activity was lacking. School B’s PE teacher proposed building a gymnasium equipped with shower facilities for the students to encourage regular and organised PA. School B’s PE teacher, who has a degree in PE, also surveys teachers about the sports activities they would be able and willing to conduct; and this allows her to organise activities that suit the school’s capability and budget. On her own initiative she also created a student health form, consisting of a student’s health status and their parents’ agreement for them to do physical activity at the school. Currently, a degree in PE is not an essential requirement for employment as a PE teacher. Based on my observations, the principal and teachers make use of their creativity to create games and play area, with minimal budgetary allowances.

## DISCUSSIONS

The use of a modified PSEA audit tool, has successfully enabled the successful identification of the various elements of primary schools in Brunei that may have contributed to an obesogenic environment. In terms of the nutrition environment, there are existing rules and regulations from the Brunei Government. However, full enforcement is still lacking in some areas, particularly the Healthy Canteen Guidelines introduced in 1994. One of the main reasons for the poor implementation of the Healthy Canteen Guidelines was that the canteen operators misunderstood or misinterpreted some of the guidelines being imposed. Previous studies have shown that having meals during school hours has impact on children’s food choices and furthermore, the school meal can contribute up to 40% of children’s daily nutrient requirement (Belot & James, 2011; Carter & Swinburn, 2004; Story et al., 2006). Therefore, full enforcement of these guidelines is very important as this affects the types of food and drinks available in the schools for the children to consume. The usefulness of my model is that it provides an overview of the elements of the schools environment that need to be considered by health workers and policy makers.

Previous studies have shown that the availability and accessibility of sports facilities in the school settings has positive impact on the children’s physical activity habits (Chomitz et al., 2011; Verstraete et al., 2006). Physical activity during recess periods has also been previously reported as an opportunity to cover about 5% to 40% of daily physical activity needs among children (Mota et al., 2005; Ridgers et al., 2005). In this research, it was found that most of the schools had inadequate sports facilities and were without suitable play areas. In most cases, the sports equipment and facilities were provided by the Brunei Government. However, poor maintenance has led to prolonged damage of the sports equipment, rendering it unusable. Based on my findings, some efforts have been demonstrated by the schools to prepare special play areas for students to use, particularly during recess.

Studies have shown that such efforts help to increase the level of physical activity among students (Ridgers et al., 2007). Despite this effort, safety is a problem due to uneven and cemented grounds, thus play requires supervision throughout the recess period. Some of the stakeholders recommended that formalisation of particular physical activity rules and regulations. There are also some useful strategies that can promote safety for students walking or cycling to school. For example, the provision of proper pedestrian and bike paths which connect the school to the housing areas. Other improvements include providing patrolled (traffic police) pedestrian crossings and strengthening road safety rules around the school area, particularly warning drivers to slow down and imposing maximum penalties.

The availability of food hawkers outside the school gate is also one of the main issues for primary schools in Brunei. The Brunei Government needs to act immediately to reduce further expansion of these vendors who currently sell mainly unhealthy food and drinks. So far there have been no formal guidelines to address this issue in Brunei. Previous studies have shown that when students pass food vendors or outlets on their way to school it undermines the messages they are given about nutrition (Walton et al., 2009). In this case, there was concern that if certain kinds of foods are being restricted within the school environment, children can still obtain these kinds of foods from the surrounding food vendors.

## CONCLUSIONS

One of the limitations of the case a study is that, although settings were selected based on their ability to provide maximum variety, it is not possible to make claims regarding the representativeness of the data for primary school settings in Brunei. Therefore, the research approach in this study was to not only examine a school’s internal environment but its external environment as well. This ensured a more complete overview of what was in-place, allowing assessment of what could be better improved to make it more conducive to childhood obesity prevention.

## Data Availability

available from the first author, SRA

## Acknowledgement

We would like to thank the Ministry of Education, Brunei Darussalam for granting permission to conduct this research, as well as all to the schools involved in this study.

## Availability of data and materials

Available by request from the corresponding author, SRA.

## Author’s Contribution

SRA and RB conceived the study question. SRA and RB drafted the study protocol, SRA completed all data analyses with RB overseeing all analyses. All authors contributed to interpretation of the findings. SRA prepared a first draft of the manuscript with input from all authors. All authors read, critically reviewed, edited, and approved the final manuscript

## Ethical Approval

Ethical approval was obtained from The University of Queensland Human Ethics Committee (Project code: 2012000915). All authors hereby declare that all experiments have been examined and approved by the appropriate ethics committee and have therefore been performed in accordance with the ethical standards laid down in the 1964 Declaration of Helsinki. Approval was obtained from the main gatekeeper, the Director of School, Ministry of Education and Ministry of Health, Brunei.

## REFERENCES

Ahmad, Siti Rohaiza (2016). Use of an audit tool to assess obesogenicity: lessons learnt from primary school environments in Brunei Darussalam. Malaysian Journal of Nutrition 2016 Vol. 22 No. 2 pp. 307-315. Doi

Bannon, K., & Schwartz, M. B. (2006). Impact of nutrition messages on children’s food choice: Pilot study. Appetite, 46(2), 124–129. doi: 10.1016/j.appet.2005.10.009

Belot, M., & James, J. (2011). Healthy school meals and educational outcomes. Journal of Health Economics, 30(3), 489–504. doi: 10.1016/j.jhealeco.2011.02.003

Birch, L. L., & Davison, K. K. (2001). Family environmental factors influencing the developing behavioral controls of food intake and childhood overweight. Pediatric Clinic North America, 48(4), 893–907. doi: 10.1016/S0031-3955(05)70347-3

Carlson, J. A., Crespo, N. C., Sallis, J. F., Patterson, R. E., & Elder, J. P. (2012). Dietary-related and physical activity-related predictors of obesity in children: a 2-year prospective study. Childhood Obesity, 8(2), 110–115. doi: 10.1089/chi.2011.0071

Carter, M. A., & Swinburn, B (2004). Measuring the ’obesogenic’ food environment in New Zealand primary schools. Health Promotion International, 19(1), 15–20. doi: 10.1093/heapro/dah103

Collingridge, D. S., & Gantt, E. E. (2008). The Quality of Qualitative Research. American Journal of Medical Quality, 23(5), 389–395. doi: 10.1177/1062860608320646

Glass, T. A., & McAtee, M. J. (2006). Behavioral science at the crossroads in public health: extending horizons, envisioning the future. Social Science & Medicine, 62(7), 1650–1671. doi: 10.1016/j.socscimed.2005.08.044

Hay, G. H. D. (2005). Normal Development in middle childhood. Psychiatry, 4(6), 3–5. doi: 10.1383/psyt.4.6.3.66355

Huston, N. R. (2006). Development Context in Middle Childhood. New York City: Cambridge University Press.

Karnik, S., & Kanekar, A. (2015). Childhood obesity: a global public health crisis. International Journal of Preventive Medicine, 3(1): 1–7. Retrieved from http://www.ncbi.nlm.nih.gov/pmc/articles/PMC3278864/

Kirk, S. F. L., Penney, T. L., & McHugh, T. L. F. (2010). Characterizing the obesogenic environment: the state of the evidence with directions for future research. Obesity Reviews, 11(2), 109–117. doi: 10.1111/j.1467-789X.2009.00611

Kral, T. V., & Rauh, E. M. (2010). Eating behaviors of children in the context of their family environment. Physiology & Behavior, 100(5), 567–573. doi: S0031-9384(10)00189-7 [pii]

Lloyd, L. J., Langley-Evans, S. C., & McMullen, S. (2012). Childhood obesity and risk of the adult metabolic syndrome: a systematic review. International Journal of Obesity (2005), 36(1), 1–11. doi: 10.1038/ijo.2011.186

Lobstein, T., Baur, L., & Uauy, R. (2004). Obesity in children and young people: a crisis in public health. Obesity Reviews, 5. doi: 10.1111/j.1467-789X.2004.00133.x

Ministry of Education. (2014). Education System: Primary Education. Retrieved from http://www.moe.gov.bn/SitePages/Primary%20Education.aspx

Ministry of Health, Brunei (2013). Health Information Booklet 2013. Bandar Seri Begawan: Retrieved from http://www.moh.gov.bn/SiteCollectionDocuments/Health%20Indicator%20Booklet/HIB_2013.pdf

Ministry of Health, Brunei (2011). Health Promotion Blueprint 2011-2015: Ministry of Health Brunei Darussalam. Bandar Seri Begawan: Retrieved from www.moh.gov.bn.

Mota, J., Silva, P., Santos, M. P., Ribeiro, J. C., Oliveira, J., & Duarte, J. A. (2005). Physical activity and school recess time: differences between the sexes and the relationship between children’s playground physical activity and habitual physical activity. Journal of Sports Science, 23(3), 269–275. doi: 10.1080/02640410410001730124

Ng, M., Fleming, T., Robinson, M., Thomson, B., Graetz, N., Margono, C., … Abera, S. F. (2014). Global, regional, and national prevalence of overweight and obesity in children and adults during 1980–2013: a systematic analysis for the Global Burden of Disease Study 2013. The Lancet, 384(9945), 766–781. doi: 10.1016/S0140-6736(14)60460-8

Ridgers, N. D., Stratton, G., & Fairclough, S. J. (2005). Assessing physical activity during recess using accelerometry. Preventive Medicine, 41(1), 102–107. doi: S0091-7435(04)00546-8 [pii]

Ridgers, N. D., Stratton, G., Fairclough, S. J., & Twisk, J. W. (2007). Long-term effects of a playground markings and physical structures on children’s recess physical activity levels. Preventive Medicine, 44(5), 393–397. doi: S0091-7435(07)00036-9 [pii]

Scaglioni, S., Arrizza, C., Vecchi, F., & Tedeschi, S. (2011). Determinants of children’s eating behavior. The American journal of Clinical Nutrition, 94(6 Suppl), 2006S–2011S. doi: 10.3945/ajcn.110.001685

Selassie, M., & Sinha, A. C. (2011). The epidemiology and aetiology of obesity: a global challenge. Best Practice and Research in Clinical Anaesthesiology, 25(1), 1–9. doi: 10.1016/j.bpa.2011.01.002

Story, M., Kaphingst, K. M., & French, S. (2006). The role of schools in obesity prevention. Future Child, 16(1), 109–142. Retrieved from http://www.ncbi.nlm.nih.gov/pubmed/16532661

Walton, M., Pearce, J., & Day, P. (2009). Examining the interaction between food outlets and outdoor food advertisements with primary school food environments. Health & Place, 15(3), 841–848. doi: 10.1016/j.healthplace.2009.02.003

Withrow, D., & Alter, D. A. (2011). The economic burden of obesity worldwide: a systematic review of the direct costs of obesity. Obesity Reviews, 12(2), 131–141. doi: 10.1111/j.1467-789X.2009.00712.x

World Health Organization. (2013). Food and Agriculture Organization of the United Nations. Diet, nutrition and the prevention of chronic diseases. Geneva: World Health Organization; 2003. WHO Technical Report Series, 916. Retrieved from http://www.who.int/dietphysicalactivity/publications/trs916/en/

